# Community-based surveys for *Plasmodium falciparum pfhrp2* and *pfhrp3* gene deletions in selected regions of mainland Tanzania

**DOI:** 10.1101/2020.05.12.20097766

**Authors:** Catherine Bakari, Sophie Jones, Gireesh Subramaniam, Celine I. Mandara, Mercy G. Chiduo, Susan Rumisha, Frank Chacky, Fabrizio Molteni, Renata Mandike, Sigsbert Mkude, Ritha Njau, Camelia Herman, Douglas P. Nace, Ally Mohamed, Venkatachalam Udhayakumar, Caleb K. Kibet, Steven G. Nyanjom, Eric Rogier, Deus S. Ishengoma

## Abstract

**Background:** Despite recent reports of false negative results among histidine-rich protein 2 (HRP2) based-malaria rapid diagnostic tests (mRDTs) caused by *pfhrp2/3* gene deletions in different countries, there is paucity of data in Tanzania.

**Methods:** This study assessed the status of *pfhrp2/3* deletions in 7,543 blood samples using laboratory multiplex antigen detection *(Plasmodium* lactate dehydrogenase - pLDH, aldolase, and HRP2). Samples showing mRDT false negativity or aberrant relationship of HRP2 to pan-*Plasmodium* antigens were genotyped for *pfhrp2/3*genes.

**Results:** Of all samples, 2,417 (32.0%) were positive for any *Plasmodium* antigens while 5,126 (68.0%) were negative. About 99.8% (n=2,411) of antigen positive samples had HRP2, but 6 (0.2%) had only pLDH and/or pAldolase. Thirteen samples had atypical relationships between pan-*Plasmodium* antigens and HRP2, but were positive by PCR. An additional 16 samples with negative HRP2 mRDTs but positive by microscopy were also chosen; all giving 35 samples genotyped for *pfhrp2/3*. Of 35 samples, 4 (11.4%) failed to consistently amplify positive control genes *(pfmsp1* and *pfmsp2)*, and *pfhrp2* and *pfhrp3* genes were successfully amplified in 31 (88.6%) samples.

**Conclusions:** Lack of *pfhrp2* and/or *pfhrp3* genes deletions in *Plasmodium falciparum* parasites supports continued use of HRP2-based mRDTs for routine malaria diagnosis in Tanzania.

## INTRODUCTION

Upon successful establishment of blood-stage infection by the *Plasmodium* parasites, various parasite proteins are produced and released into the host blood. Some of these proteins (also referred to as antigens) are targets for malaria rapid diagnostic tests (mRDTs). Three antigen targets currently in use include the *Plasmodium* lactate dehydrogenase (pLDH), *Plasmodium* aldolase (pAldolase), and the *Plasmodium falciparum-specific* histidine rich protein 2 (HRP2) [1–3]. The use of antigen-based mRDTs in many malaria endemic countries worldwide have profoundly improved malaria case management and surveillance efforts and remains an essential diagnostic tool, especially in Africa [4–7].

HRP2-based tests are species-specific since the antigen is only produced by *P falciparum*, though tests detecting pLDH and pAldolase have the potential to detect all human malarias [2,4,8]. HRP2 is the most widely used antigen in mRDTs either alone or in combination with other antigens, due to its abundance, specificity for *P. falciparum* infection, and high sensitivity and thermal stability [9]. Antibodies raised against HRP2 can cross-react with the *P. falciparum* HRP3 antigen due to similarities in amino acid sequences and repeating epitopes [10–12]. The genes encoding for these two antigens are located on different chromosomes of the *P. falciparum* genome, with *pfhrp2* on chromosome 8 while *pfhrp3* gene is on chromosome 13 [13,14]. Large number of genetic deletions of *pfhrp2* and/or *pfhrp3* genes in natural populations of *P. falciparum* were first reported in parasite populations in Peru and subsequently in many countries including Africa [15–22].

Sensitivity of HRP2-based mRDTs can be affected by transportation and storage conditions outside of manufacturer specifications, operator errors, low density infections, or a mutation or deletion of the *pfhrp2* and/or *pfhrp3* genes in the infecting parasite strain [15–23]. In addition, the diversity in parasite population and number of epitopes on HRP2 recognized by the diagnostic test antibodies may modify the sensitivity of the test when dealing with different *P. falciparum* populations [24–28].

In Tanzania, mRDTs were introduced between 2009 and 2012 and are now widely used in both private and public health facilities throughout the country. Before the introduction of mRDTs in Tanzania, a study conducted between 2005 and 2010 (the African Quinine Artesunate Malaria Treatment Trial) found no evidence of *pfhrp2/3* gene deletions [29]. However, *pfhrp2* gene deletions have been reported in the neighbouring East African countries, including Kenya [21] and Rwanda [27]. In addition, a study conducted in Tanzania which analysed samples collected in 2010 showed evidence of sporadic occurrence of *pfhrp2/3* gene deletions in some areas [30]. Although both the initial evidence (sample confirmed as microscopy positive *for P. falciparum* but negative PfHRP2-detecting mRDTs) and confirmatory evidence (molecular approaches) [31] were used to screen for *pfhrp2/3* gene deletions, the sample size and the geographic regions covered were limited. In this study, field diagnostic results and a multiplex antigen detection assay was used to investigate potential *pfhrp*2/3 gene deletions in samples collected in 2017 from four regions of Tanzania with persistently high malaria transmission after five years of introduction of mRDTs.

## MATERIALS AND METHODS

### Study sites

Samples and data were obtained from a cross-sectional survey conducted between July and November 2017 in four regions of Tanzania (Geita, Kigoma, Mtwara and Ruvuma). These regions were among those with persistently high malaria transmission as shown by the surveys conducted from 2007 – 2017 [32–35]. The four Regions also had higher prevalence in the School Malaria Parasitological Survey (SMPS) of 2014/2015 [36], and are among the 10 regions targeted by the National Malaria Control Programme (NMCP) for reduction of malaria burden through the high burden to high impact initiative (WHO and NMCP revised strategic plan). Two districts with high prevalence in the SMPS of 2014/2015 were purposively selected from each region; Nyang’hwale and Chato (Geita), Buhigwe and Uvinza (Kigoma), Mtwara DC and Nanyumbu (Mtwara) and Nyasa and Tunduru (Ruvuma). Within each district, two villages were purposively selected for sampling based on the malaria parasite positivity rates as reported from health facility reports, making a total of 16 villages sampled all members of the selected households (HHs) were asked to participate in the survey [37]. Blood samples were collected by finger prick, thin and thick films were prepared, and all study participants were screened with malaria mRDT as per the manufacturer’s instructions. CareStart Malaria HRP2/pLDH (Pf/PAN) COMBO (AccessBio,NJ, USA) mRDTs were used in Geita and Kigoma regions, and Lundo village of Nyasa District (Ruvuma region) while CareStart Malaria HRP2 (Pf) (AccessBio,NJ, USA) were used in the rest of the villages in Ruvuma and Mtwara regions. The results were interpreted within the specified reading time of the manufacturer’s protocol. Dried blood spots on filter papers (DBS) were collected on Whatman 3MM paper (GE Healthcare, PA, USA), dried for 2-4 hours, and individually packaged in sealable plastic bags with desiccant for further laboratory analysis. Participants with mRDT positive results were treated according to the national guidelines [38].

The main study and the laboratory analyses reported in this manuscript obtained ethical approval from the Medical Research Coordinating Committee (MRCC) of the National Institute for Medical Research (NIMR), and permission to conduct the study in the selected regions was sought from the President’s Office, Regional Administration and Local Government Authority and Regional, District and village authorities. Informed consent/assent was sought before conducting the demographic survey or including the participants into the study. The laboratory activities undertaken at CDC were considered non-research by the CDC Human Subjects office for the purpose of providing laboratory testing of these specimens and participation of CDC scientists for this collaboration.

### Microscopy

Thick and thin blood films for parasite counting and species identification were prepared from the finger prick blood and stained using 2.5–3% Giemsa for 45–60 minutes. The films were examined by trained microscopists to detect parasite infection status and parasite density using thick films while parasite species were assessed on thin films. Parasites were counted as asexual parasites per 200 White Blood Cells (WBCs) for asexual parasites or 500 WBCs for sexual stages. A blood film was declared negative if no *Plasmodium* parasites were detected after examining 200 high power fields for the thick film. Parasite density (parasites per μL of blood) was calculated by multiplying the number of asexual parasites by 40 or sexual stages by 16 assuming one microliter of blood contained 8000 WBC.

### Sample processing and Laboratory Multiplex Assay

DBS were shipped to the Malaria Laboratory, Centers for Disease Control and Prevention, Atlanta, under ambient temperature. A 6mm punch of each sample was taken and eluted in blocking buffer containing: PBS, 0.5% polyvinyl alcohol (Sigma, St. Louis, MO), 0.8% polyvinylpyrrolidine (Sigma), 0.1% casein (ThermoFisher Scientific, Waltham, MA), 0.5% BSA (Sigma), 0.3% Tween20, 0.05% sodium azide, and 0.01% *E. coli* extract to prevent non-specific binding. The elution step diluted the samples to a 1:20x whole blood dilution, which was the dilution used for the assay. DBS samples were screened by a bead-based multiplex antigen assay for the simultaneous detection of *P. falciparum* HRP2 (HRP2), pan-*Plasmodium* aldolase (pAldolase), and pan-*Plasmodium* lactate dehydrogenase (pLDH) by previously-described protocol [39]. Antibodies used to detect epitopes on HRP2 would also bind to the same epitopes on the HRP3 antigen. Protocol details are outlined in the Supplemental Data.

### DNA Extraction, PET-PCR, *pfhrp2* and *pfhrp3* genotyping

For samples selected for further molecular characterization, genomic DNA was extracted from DBS using the QIAamp DNA Mini Kit (Hilden, Germany) using manufacturers protocol and screened for parasite DNA using the multiplex photo-induced electron transfer PCR (PET-PCR) assay as previously described [40–43]. PCR for *pfhrp2* and *pfhrp3* genotyping was performed as described previously [44,45]. Complete details for the molecular assays are outlined in Supplemental Data: (Supplemental Table1; Details of Primers and PCR reaction conditions to amplify *pfhrp2* and *pfhrp3* genes and Supplemental Table 2; Details of Primers and PCR reaction conditions to amplify *msp-1* and *msp-2* genes).

### Data analysis

The database and the different data collection applications were created using the Open Data Kit (ODK) software. Data cleaning, validation and quality control were undertaken as described by Chiduo *et al* [37].The data was later transferred to Microsoft Excel (Redmond, WA, USA) and STATA software (Texas, USA) used for analysis which involved generating a summary of basic features of the study population.

To determine if a sample’s laboratory mean fluorescence intensity minus background signal (MFI-bg) was to be denoted as positive for a specific antigen, two methods were employed. First, a panel of 24 known negative blood samples which had been eluted from Whatman 903 filter paper were run by the multiplex antigen assay, the lognormal mean and standard deviation was derived from this sample set. The mean +3sd was calculated to provide a MFI-bg threshold signal which was used as a cutoff to define antigen positive samples. Second, a two-component finite mixture model was used for the log-transformed antigen MFI-bg data from the study, and the mean +3sd of the first component was used to define this cutoff. In order to reduce Type I errors, the more conservative of these two methods was used to determine the MFI-bg signal where any sample values above this would be considered a true positive for that particular antigen [46].

In comparing HRP2 antigen signal to either of the pan*-Plasmodium* markers, the typical relationship between these two antigens was defined as the standard correlation observed for the vast majority of the observations. Visual outliers to this standard correlation were identified as outliers with suspicion of aberrant HRP2 production by the *P*. *falciparum* parasite requiring further molecular investigation.

## RESULTS

A total of 2,520 households with 7,313 individuals were sampled for the cross-sectional survey, which was conducted in 16 villages (in 8 districts) from four regions of Tanzania (Geita, Kigoma, Mtwara and Ruvuma) between July and November 2017 (Table 1). Apart from the 7,313 individuals enrolled, an additional of 230 samples were taken from individuals who came to seek health services, but were not among the 120 sampled households. Therefore, 7,543 blood samples were available for this study, and 3.0% (230/7,543) of these were from individuals with incomplete data. The remainder (97.0%, 7,313/7,543) had complete data with parasitological, clinical and demographic information. The mean age in years was 22.3 (SD=21.0), and 43.4% of participants were male (Table 1).

**Table 1:**
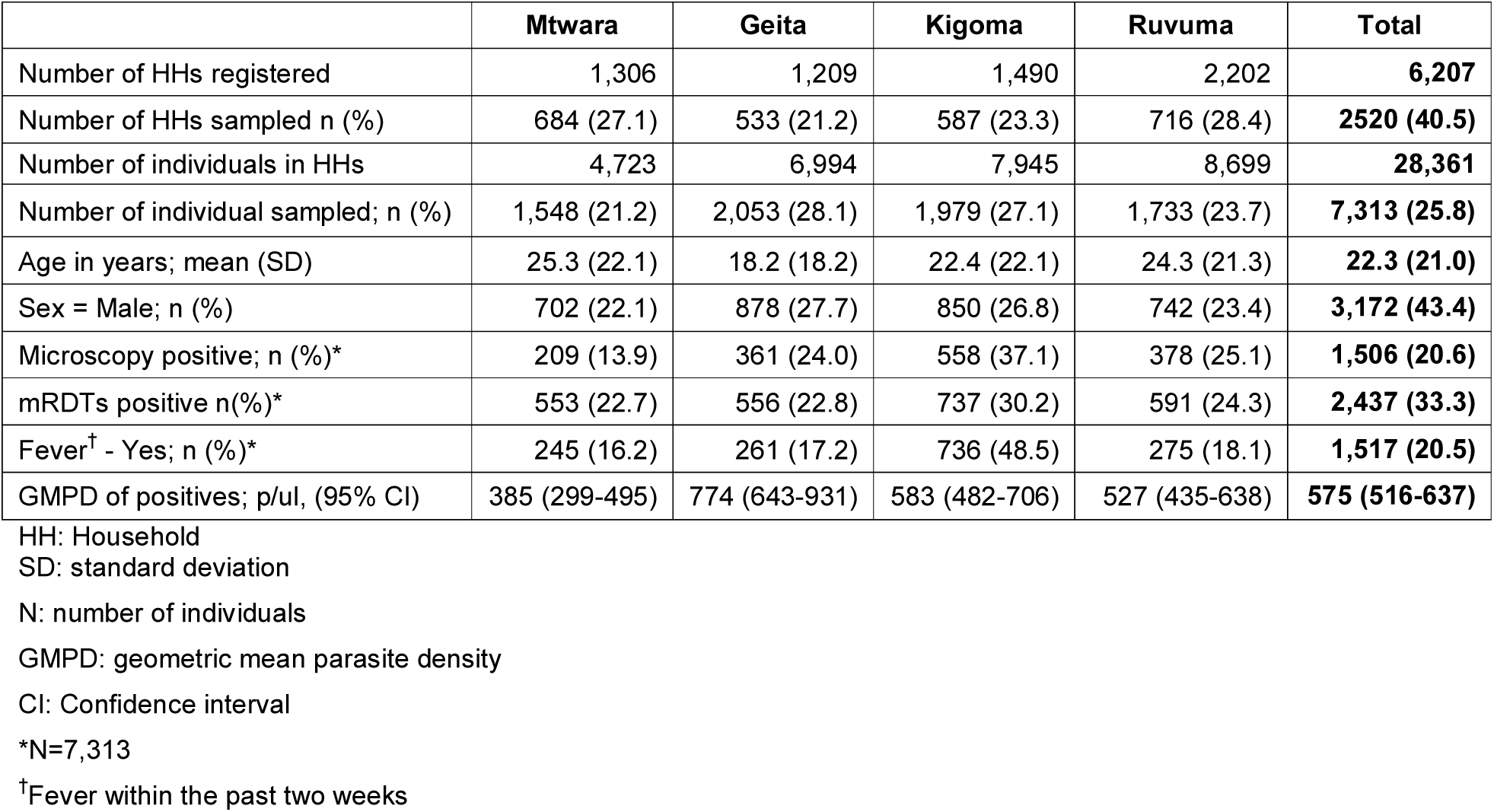
Baseline characteristics of individuals sampled in each of the four regions

For all persons enrolled in all study sites, 38.4% (2897/7543) were positive for any one of the three parasite detection assays: microscopy, mRDT, or bead-based multiplex assay. The results showed 15.8% (1,158/7,313) of the persons were positive for *P. falciparum* infection by microscopy, 33.3% (2,437/7,313) were positive by HRP2 mRDT, and 32.0% (2,417/7,543) were positive for any one of the *Plasmodium* antigens tested by the bead-based multiplex assay. Concordance among these three malaria tests is shown in Supplemental Figure 1. For those positive by microscopy, the geometric mean parasite density was 575 asexual parasites/μL of blood.

**Figure 1:**
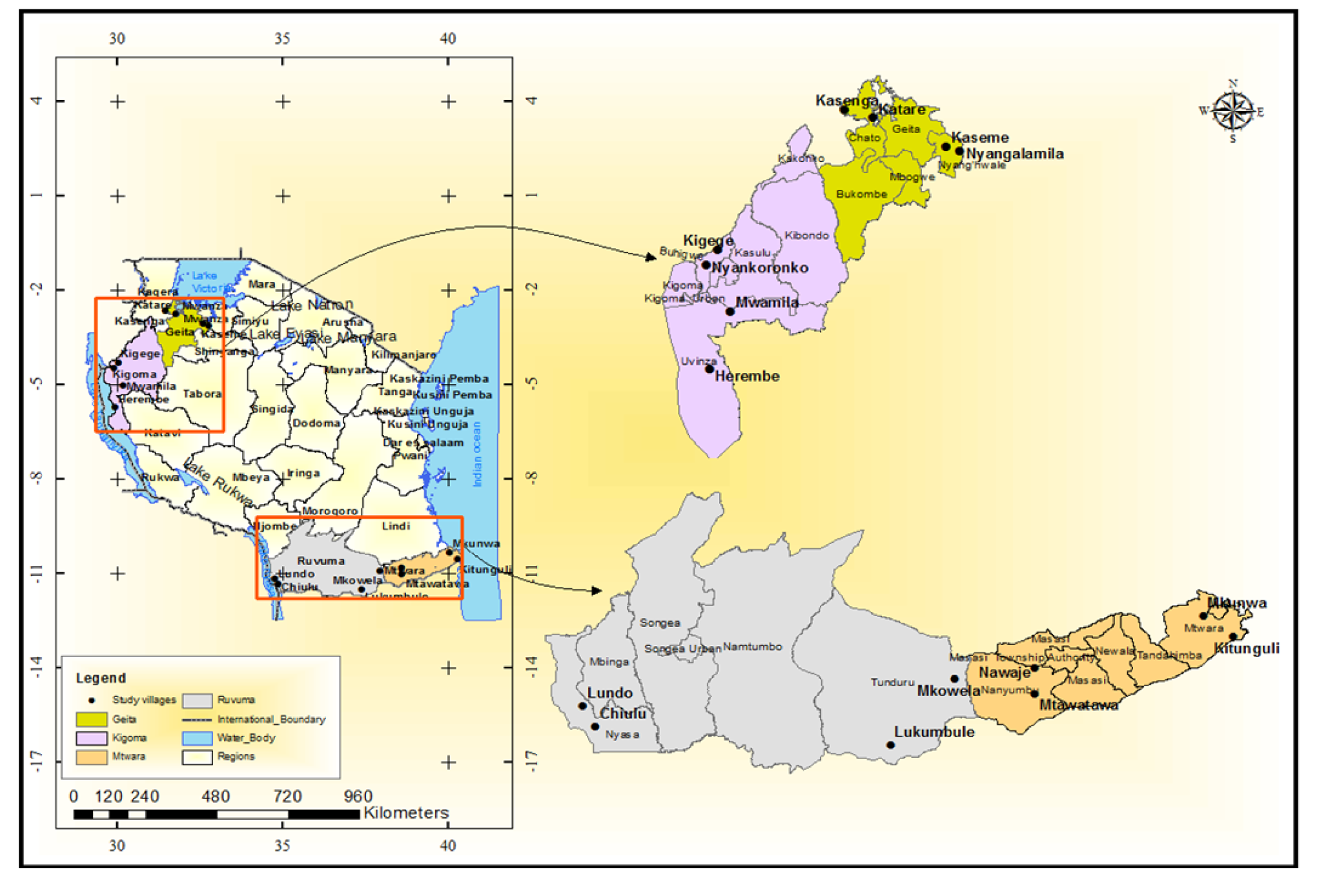
Map showing the study sites in the four regions of Tanzania

A flow diagram for sample selection for further molecular testing to detect the presence (and potential prevalence) of *pfhrp2* and *pfhrp3* deletions in this study population is shown in Figure 2. In selecting specimens warranting molecular assays for *pfhrp2* and *pfhrp3* genotyping, two types of information were considered: discordance between field microscopy and mRDT results for an individual, and relationship between the pan*-Plasmodium* antigens and the HRP2 antigen for an individual’s blood sample. From the field tests (microscopy and mRDT), 95 persons were found to be microscopy positive but mRDT negative (1.3% of the 7313 persons who had data for both tests). Of these 95 persons, 94 had a DBS available for multiplex antigen detection, and 54.3% (51/94) of these were found to be HRP2 antigen positive by the laboratory test with a typical relationship to the other pan-*Plasmodium* antigens (as illustrated in Figure 3). Additionally, 28.7% (27/94) of these samples were negative for *P. falciparum* DNA and could not undergo genotyping. The remaining 16 samples (17.0% of the 94 selected by microscopy/mRDT discordance) were all *P. falciparum* DNA positive, but had no antigens detected (n=7) or an atypical relationship between the pan-*Plasmodium* markers and HRP2 (n=9). Based on these test results, these 16 were selected as warranting *pfhrp2* and *pfhrp3* genotyping.

**Figure 2.**
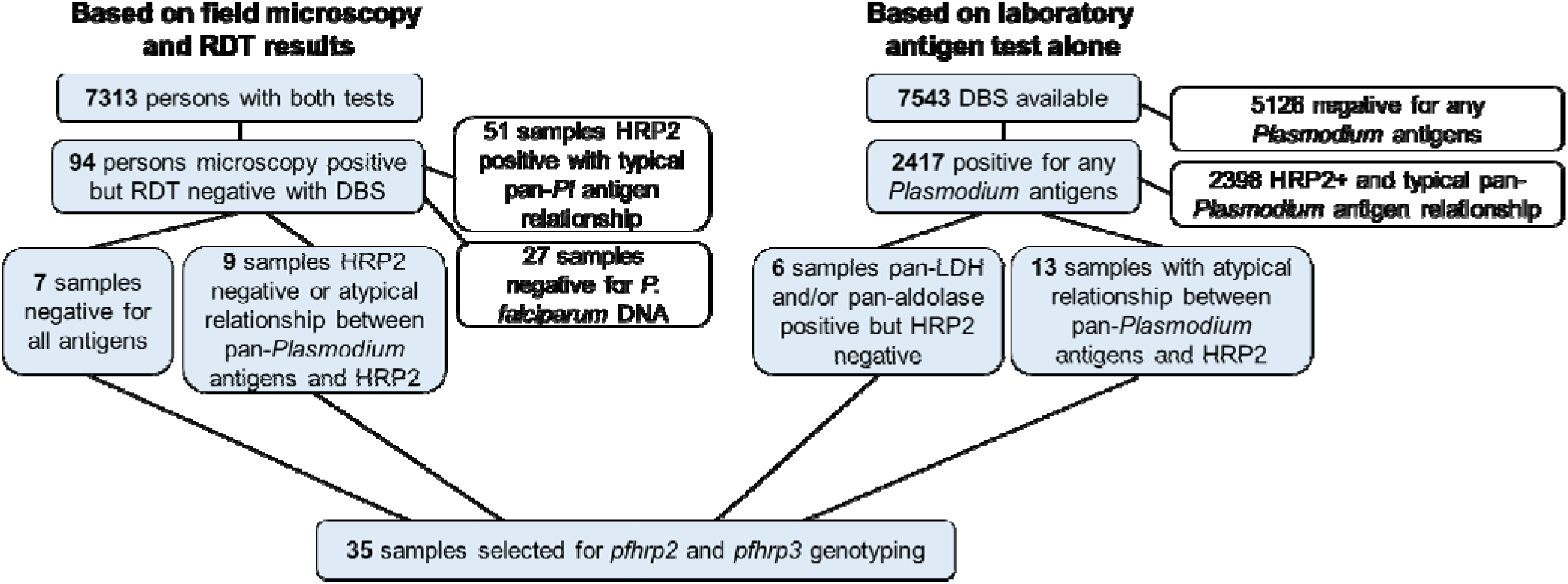
Flowchart for selection of samples requiring genotyping for *pfhrp2* and *pfhrp3*. Samples were selected based on initial field microscopy and mRDT results (shown on left), or multiplex laboratory antigen detection (shown on right), for final determination of samples requiring genotyping to detect potential deletion of *pfhrp2/3* genes.

Since all DBS were screened by the multiplex antigen assay, in addition to samples selected for genotyping by the field diagnostic tests, samples could be selected for *pfhrp2* and *pfhrp3* genotyping based solely on laboratory results. Of all 7,543 DBS screened by the multiplex antigen assay, malaria antigen could not be detected in 5,126 (68.0%) samples. Of the 2,417 DBS positive for any antigens, 2,398 (99.2%) of these were found to have a typical relationship of the pan-*Plasmodium* markers with HRP2 (Figure 3). Of the remaining 19 DBS which were all positive for the pan-*Plasmodium* antigens; 6 had a complete absence of HRP2 antigen, and 13 had an atypical relationship between the assay signal for the pan marker and HRP2. All of these 19 were positive for *P. falciparum* DNA and could thus be utilized for genotyping PCRs. Together with the 16 samples chosen initially based on field test results, these 35 samples (16 + 19) were gathered as the final set with suspicion of aberrant HRP2 and/or HRP3 antigen production and warranted genotyping for the *pfhrp2* and *pfhrp3* genes.

**Figure 3.**
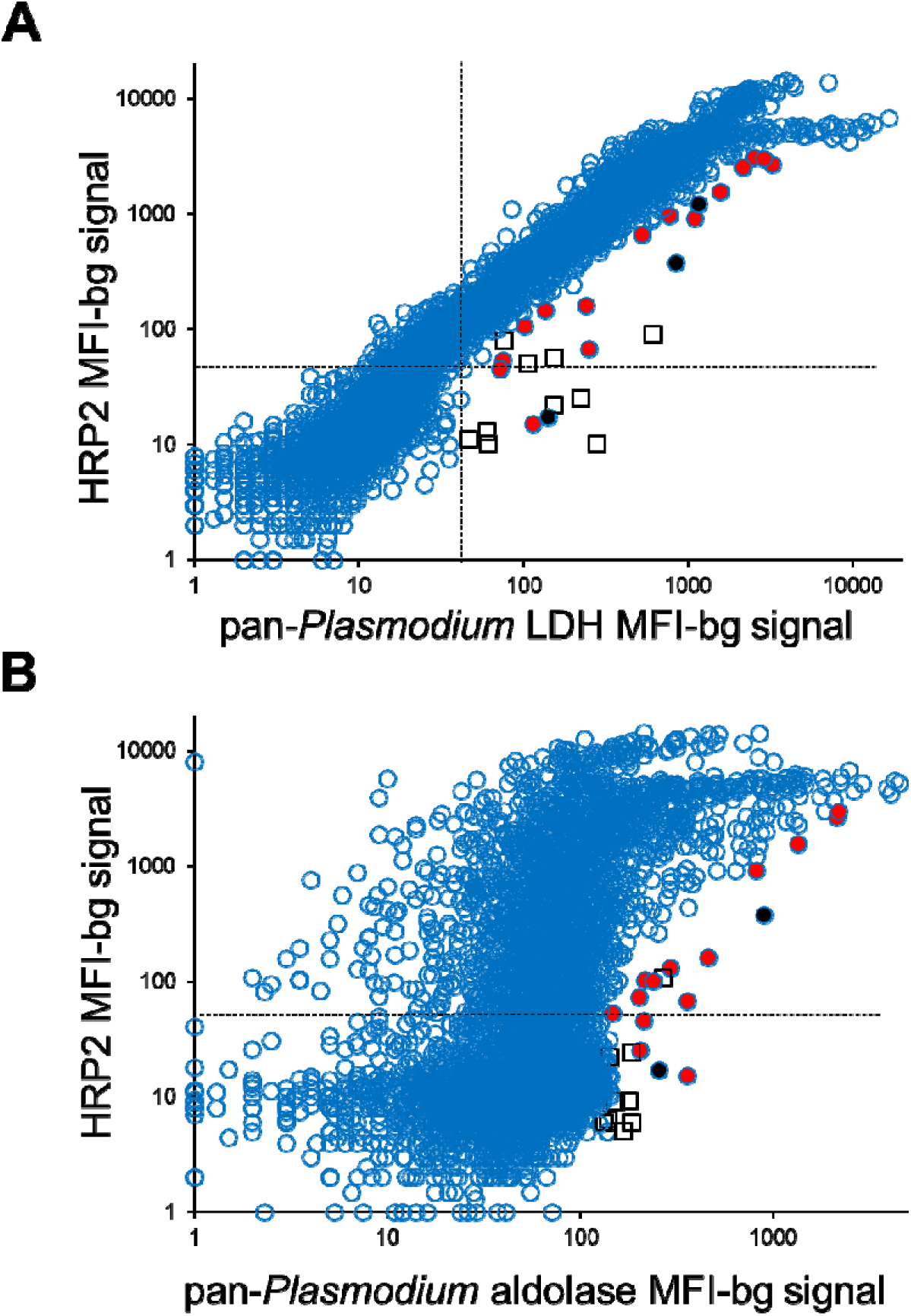
Scatterplots of pan*-Plasmodium* LDH or aldolase assay signal in comparison with HRP2 assay signal. Plots designate samples that were selected for further genotyping investigation as based on an atypical relationship to the pan*-Plasmodium* LDH (A) or aldolase (B) antigens. Hashed lines in each plot show MFI-bg assay signal threshold which would indicate a positive assay signal for each antigen. Black circles indicate samples selected based on field RDT result as well as laboratory antigen assay that were *P falciparum* DNA positive. Red circles indicate samples selected solely based on laboratory antigen assay that were *P. falciparum* DNA positive. Squares i indicate samples selected solely based on laboratory antigen assay that were *P. falciparum* DNA negative.

Table 2 outlines the *pfhrp2* and *pfhrp3* genotyping results for these 35 samples, as well as other information regarding characteristics of the individuals, field test results, and other factors. Most of the samples (31/35, 88.6%) were found to successfully amplify the *pfhrp2* and *pfhrp3* genes for the two exons of each gene. However, one or more of the *pfhrp2* and *pfhrp3* exon targets could not be amplified for 4 (11.4%) DNA samples. To correctly report the presence of a deletion (i.e. lack of PCR amplification), we chose to include amplification targets for two other single-copy genes (*pfmspl* and *pfmsp2)* to verify true non-amplification event [31]. For these 4 DNA samples, all were unable to consistently amplify both *pfmsp1* and *pfmsp2* single-copy genes and were excluded in the analysis. For this reason, non-amplification of *pfhrp2* and *pfhrp3* targets due to true deletion events could not be verified, and thus, no deletions in these genes could be confirmed in the remaining 31 samples.

**Table 2.**
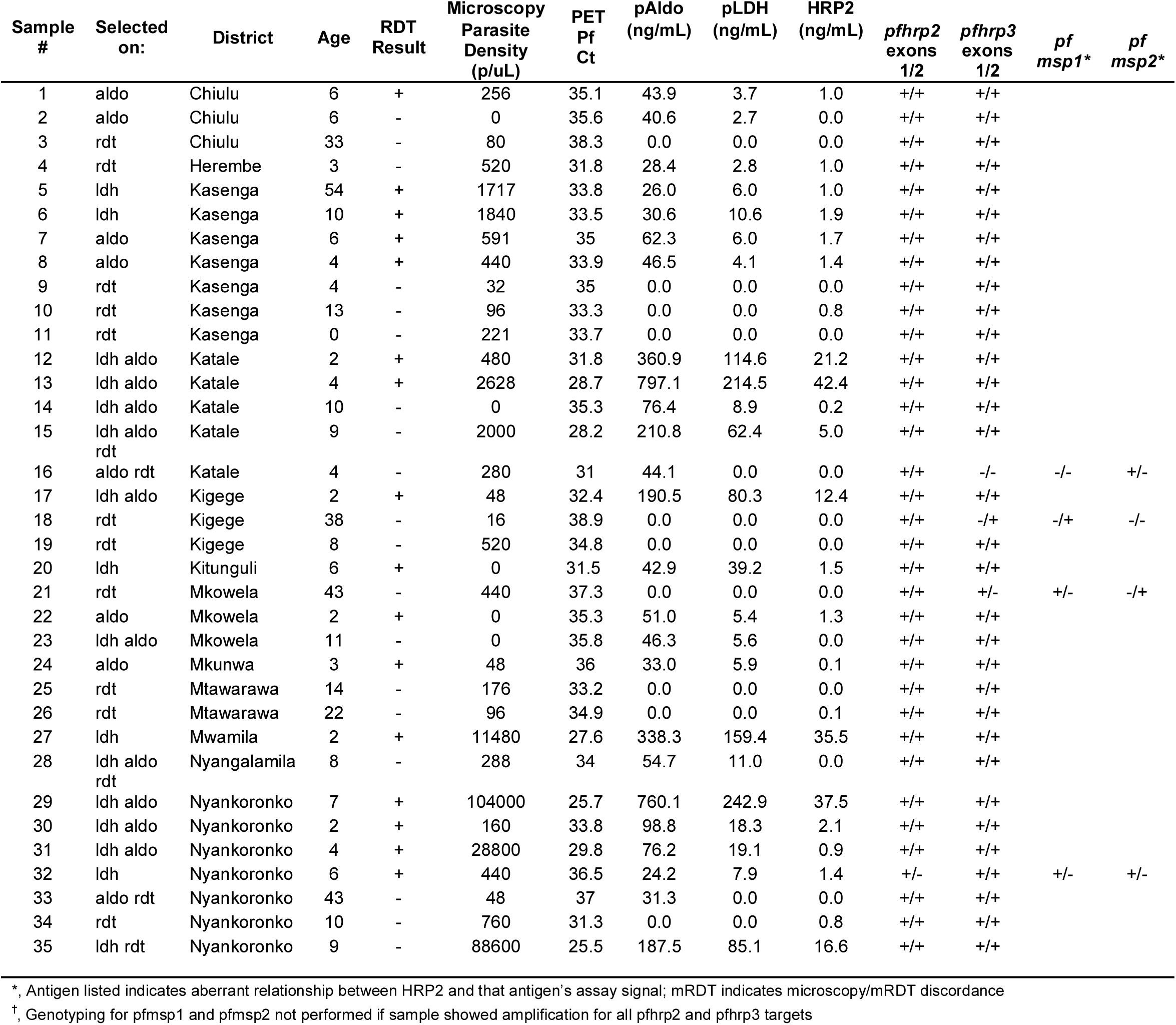
Summary of *pfhrp2* and *pfhrp3* genotyping results for the samples selected based on field mRDT and microscopy results, laboratory antigen assay, or both.

## DISCUSSION

The samples used in this study were collected during a community-based survey which was conducted in four regions with persistently high malaria burden over the past 10yrs and used to assess the presence and prevalence of *pfhrp2* and *pfhrp*3 gene deletions in Tanzania. The findings from field mRDT and microscopy tests as well as the laboratory multiplex antigen test indicate that the vast majority of *P. falciparum* infections in Tanzania produced high levels of HRP2 (and HRP3) antigens which would be recognized by HRP2-based mRDTs. Although mRDTs did not detect some infections from persons confirmed to be *P. falciparum* positive by PCR, the evidence presented here suggests that these false negative mRDT results were not due to *pfhrp2* and *pfhrp3* gene deletion, and more a factor of low parasite density infections.

During the survey, extensive efforts were taken to protect the quality of the mRDTs. The experienced study team ensured that mRDTs were stored in appropriate conditions as per the manufacturers’ instructions; transported in good conditions (ambient temperature with minimal humidity) and the tests were also performed by experienced technician. To minimize operator errors, the testing process and reading of mRDT results were done in the presence of other members of the team who ensured that any doubtful mRDTs results were correctly read and interpreted. Studies conducted elsewhere reported that the performance of HRP2-based mRDTs depends on the level of parasitaemia [1,8,47], with the lower limit of detection generally around 200 parasites/μL[1]. This community survey included mainly asymptomatic persons, and some had low-level (sub-detectable by mRDT) parasitaemia; this could possibly explain some of the discordance between the field mRDT results and the laboratory antigen test. In total 1,998 (27.3% of all) persons were concordant between those two tests, whereas 372 (6.0%) individuals were positive for the mRDT alone and 325 (4.4%) were positive only by the laboratory antigen test. As mRDTs are designed for reliable detection of parasite densities more typical of clinical relevance (200p/uL or greater), their use in the asymptomatic population may miss low-density infections and HRP2 levels [46]. Even in this community setting, it was encouraging to see that mRDTs were able to detect the majority of persons with malaria antigenemia as determined by the laboratory antigen test. Of 2,323 total samples found to be antigen positive, 1,998 (86.0%) came from persons who were mRDT positive. Considering the generally low parasite densities for any *P. falciparum*-infected persons in this survey, concordance was also good for all three tests; microscopy, RDTs, and the laboratory antigen test. Of 1,158 total microscopy positives, 1,063 (91.8%) were also mRDT positive, and 1,050 (90.7%) were also positive by the laboratory antigen test. Concordance among multiple malaria indicators provides greater confidence for the true levels of malaria in a populace.

Of the 35 samples selected for further molecular investigation, which were positive for *P. falciparum* by PCR, the majority (27/35) had relatively low parasite densities ranging from 0 to 1,000 parasite/μl. However, eight samples were from persons with higher parasite densities (>1,000 asexual parasites/μl), and two of these samples had very high parasite density (88,600 and 104,000 parasite/μl). Two of these eight persons gave negative mRDT results which could potentially be explained by the prozone effect where excess of antigen leads to false-negative results [48]. All eight of these higher density infections were found to have detectable HRP2 antigen, though at much lower blood concentrations than typical given those levels of *P. falciparum* parasite densities.The multiplex antigen screening allowed for reconfirmation of HRP2 (and possibly HRP3) antigen profiles for DBS samples. Of all 7,543 samples screened by the multiplex antigen test, few (28, 0.4%) samples had a complete absence of HRP2 or an aberrant relationship between the assay signal for the pan marker and HRP2. This could be due to very low parasite densities or infection with a non-*P. falciparum* infection, but this observation could not be explained by deletions of the *pfhrp2* and *pfhrp3* genes.

Our report adds to the literature in the same way as studies in Honduras [45] and French Guiana

[49] where no deletion of *pfhrp2* gene were reported. However, these findings are in contrast with the results from a recent study which was conducted in Mbeya, Mtwara, and Mwanza regions in Tanzania using samples collected in 2010, one year following the introduction of mRDTs. That study found evidence of sporadic occurrence of *pfhrp2 and pfhrp3* gene deletion in some areas, with 1.7% of isolates tested reported to have a deletion of either of the genes [30]. Though recommended molecular tests [31] were used in this study, the overall study was very small with only 176 Tanzanian samples tested. Malaria endemicity and year of sampling could potentially explain the different results between the present and the previous surveys, but with the very low estimates for prevalence from the 2010 study, there is a potential that those parasite strains are still present at very low- levels in these regions.

A limitation of the current study is that single gene deletions of *pfhrp2* or *pfhrp3* alone could be masked by a parasite producing at least one of these antigens. Deletions (or loss-of-function mutations) in both genes would lead to a complete absence of HRP2/HRP3 antigens in a *P. falciparum* infection. However, if one of these antigens was being produced by the parasite, the algorithm defined here may not identify the blood sample as suspicious if the assay signal for HRP2 remains high. Additionally, the molecular tests performed here were only for complete gene deletions, and any loss-of-function point mutations leading to antigen non-expression would not be captured since the *pfhrp2* or *pfhrp3* specific DNA amplification would still occur using the current protocol. Another limitation from the study is that it does not provide estimates for entire country, since only 4 out of the 26 regions of mainland Tanzania were included in this study. Further studies would need to be conducted on *P. falciparum* isolates collected from other geographical regions of Tanzania (especially in low transmission areas) in order to increase the chances of detecting *pfhrp2* and *pfhrp3* gene deletion in different parts of the country.

## CONCLUSIONS

Though a low number of false negative mRDT results were found in Tanzania, these could not be explained by *pfhrp2* or *pfhrp3* gene deletions. Overall the study results suggest that HRP2-based mRDTs for detection of *P. falciparum* infection and confirmatory diagnosis of malaria in the surveyed area in Tanzania can be used as a reliable tool for malaria case management.

## Data Availability

The datasets generated during and/or analysed during the current study are available from the corresponding author on reasonable request

## Acknowledgments

The authors would like to thank the survey teams and study participants for their involvement in the survey in which the tested samples were collected. CB and DSI are supported by DELGEME through the DELTAS Africa Initiative (DELGEME grant 107740/Z/15/Z). The DELTAS Africa Initiative is an independent funding scheme of the African Academy of Sciences (AAS)’s Alliance for Accelerating Excellence in Science in Africa (AESA) and supported by the New Partnership for Africa’s Development Planning and Coordinating Agency (NEPAD Agency) with funding from the Wellcome Trust (DELGEME grant 107740/Z/15/Z) and the UK government. The views expressed in this publication are those of the author(s) and not necessarily those of AAS, NEPAD Agency, Wellcome Trust, or the UK government.

## Author Contributions

Coordination of field surveys: C.I.M., M.G.C., S.R., F.C., F.M., R.M., S.M., A.M., C.K.K., S.G.N., D.S.I. Conceived and designed experiments: V.U., E.R., D.S.I. Performed the experiments: C.B., S.J., G.S., C.H., D.P.N., E.R. Analyzed the data: C.B., E.R., D.S.I. Contributed reagents, materials, and analysis tools: V.U., E.R. Manuscript preparation: C.B., V.U., E.R., D.S.I. All authors reviewed and approved the final manuscript.

## Disclaimer

The findings and conclusions presented in this report are those of the authors and do not necessarily reflect the official position of the Centers for Disease Control and Prevention. Ritha Njau is a staff member of the World Health Organization. She alone is responsible for the views expressed in this publication, which do not necessarily represent the decisions, policy or views of the World Health Organization.

## Footnotes

This work was supported by The Global Fund, CDC and DELGEME Project.

The authors declare they have no competing interests. None of the material has been presented or published previously.

## REFERENCES

1. Moody A. Rapid Diagnostic Tests for Malaria Parasites. 2002; 15 (1): 66–78.

2. WHO. New Perspectives: Malaria Diagnosis: Report of a Joint WHO/USAID Informal Consultation. Who [Internet]. 2000; (October 1999):1–29. Available from: http://www.who.int/malaria/publications/atoz/who_cds_rbm_2000_14/en/%0Ahttp://www.who.int/tdr/publications/documents/malaria-diagnosis.pdf

3. Maltha J, Gillet P, Jacobs J. Malaria rapid diagnostic tests in endemic settings. Clin Microbiol Infect [Internet]. European Society of Clinical Infectious Diseases; 2013; 19(5): 399–407. Available from: http://dx.doi.org/10.1111/1469-0691.12151

4. Bell D, Wongsrichanalai C, Barnwell JW et al. Ensuring quality and access for malaria diagnosis: How can it be achieved? Nat Rev Microbiol. 2006; 4(9):682–695.

5. Masanja MI, McMorrow M, Kahigwa E, Kachur SP, McElroy PD. Health workers’ use of malaria rapid diagnostic tests (RDTS) to guide clinical decision making in rural dispensaries, Tanzania. Am J Trop Med Hyg. 2010; 83(6): 1238–1241.

6. Rutta ASM, Francis F, Mmbando BP, et al. Using community-owned resource persons to provide early diagnosis and treatment and estimate malaria burden at community level in north-eastern Tanzania. Malar J. 2012; 11: 1–8.

7. Thiam S, Thior M, Faye B, et al. Major Reduction in Anti-Malarial Drug Consumption in Senegal after Nation-Wide Introduction of Malaria Rapid Diagnostic Tests. PLoS One. 2011; 6(4).

8. Murray CK, Gasser RA, Magill AJ, Miller RS. Update on rapid diagnostic testing for malaria. Clin Microbiol Rev. 2008; 21(1): 97–110.

9. WHO. Global Malaria Programme WHO: World Malaria Report 2012. 2012;:V–195.

10. Rock EP, Marsh K, Taylor DW, et al. Comparative analysis of the Plasmodium falciparum histidine-rich proteins HRP-I, HRP-II and HRP-III in malaria parasites of diverse origin. Parasitology. 1987; 95(2):209–227.

11. Lee N, Baker J, Andrews KT, et al. Effect of sequence variation in Plasmodium falciparum histidine-rich protein 2 on binding of specific monoclonal antibodies: Implications for rapid diagnostic tests for malaria. J Clin Microbiol. 2006; 44(8):2773–2778.

12. Lee N, Gatton ML, Pelecanos A, et al. Identification of optimal epitopes for Plasmodium falciparum rapid diagnostic tests that target histidine-rich proteins 2 and 3. J Clin Microbiol. 2012; 50(4): 1397–1405.

13. Wellems TE, Howard RJ. Homologous genes encode two distinct histidine-rich proteins in a cloned isolate of Plasmodium falciparum. Proc Natl Acad Sci U S A [Internet]. 1986; 83(16): 6065–9. Available from: http://www.pubmedcentral.nih.gov/articlerender.fcgi?artid=386439&tool=pmcentrez&rendertype=abstract

14. Kemp DJ, Thompson JK, Walliker D, Corcoran LM. Molecular karyotype of Plasmodium falciparum: conserved linkage groups and expendable histidine-rich protein genes. Proc Natl Acad Sci. 2006; 84(21):7672–7676.

15. Gamboa D, Ho MF, Bendezu J, et al. A large proportion of P. falciparum isolates in the Amazon region of Peru lack pfhrp2 and pfhrp3: Implications for malaria rapid diagnostic tests. Bjorkman A, editor. PLoS One [Internet]. Public Library of Science; 2010 [cited 2018 Apr 6]; 5(1):e8091. Available from: http://dx.plos.org/10.1371/journal.pone.0008091

16. Maltha J, Gamboa D, Bendezu J, et al. Rapid Diagnostic Tests for Malaria Diagnosis in the Peruvian Amazon: Impact of pfhrp2 Gene Deletions and Cross-Reactions. PLoS One. 2012; 7(8): 1–7.

17. Koita OA, Doumbo OK, Ouattara A, et al. False-negative rapid diagnostic tests for malaria and deletion of the histidine-rich repeat region of the hrp2 gene. Am J Trop Med Hyg. 2012; 86(2): 194–198.

18. Berhane A, Russom M, Bahta I, Hagos F, Ghirmai M, Uqubay S. Rapid diagnostic tests failing to detect Plasmodium falciparum infections in Eritrea: An investigation of reported false negative RDT results. Malar J. BioMed Central; 2017; 16 (1): 1–6.

19. Parr JB, Verity R, Doctor SM, et al. Pfhrp2 -Deleted Plasmodium falciparum Parasites in the Democratic Republic of the Congo: A National Cross-sectional Survey. J Infect Dis. 2017; 216(1): 36–44.

20. Viana GMR, Okoth SA, Silva-Flannery L, et al. Histidine-rich protein 2 (pfhrp2) and pfhrp3 gene deletions in Plasmodium falciparum isolates from select sites in Brazil and Bolivia. PLoS One. 2017; 12(3):1–13.

21. Beshir KB, Sepúlveda N, Bharmal J, et al. Plasmodium falciparum parasites with histidine-rich protein 2 (pfhrp2) and pfhrp3 gene deletions in two endemic regions of Kenya. Sci Rep. 2017; 7(1): 1–10.

22. Bharti PK, Chandel HS, Ahmad A, Krishna S, Udhayakumar V, Singh N. Prevalence of pfhrp2 and/or pfhrp3 gene deletion in plasmodium falciparum population in eight highly endemic states in India. PLoS One. 2016; 11 (8): 1–16.

23. Baker J, McCarthy J, Gatton M, et al. Genetic Diversity of *Plasmodium falciparum* Histidine Rich Protein 2 (PfHRP2) and Its Effect on the Performance of PfHRP2 Based Rapid Diagnostic Tests. J Infect Dis [Internet]. 2005; 192(5):870–877. Available from: https://academic.oup.com/jid/article-lookup/doi/10.1086/432010

24. Wurtz N, Fall B, Bui K, et al. Pfhrp2 and pfhrp3 polymorphisms in Plasmodium falciparum isolates from Dakar, Senegal: Impact on rapid malaria diagnostic tests. Malar J. 2013; 12(1): 1–8.

25. Atroosh WM, Al-Mekhlafi HM, Al-Jasari A, et al. Genetic variation of pfhrp2 in Plasmodium falciparum isolates from Yemen and the performance of HRP2-based malaria rapid diagnostic test. Parasites and Vectors [Internet]. Parasites & Vectors; 2015; 8(1): 1–8. Available from: http://dx.doi.org/10.1186/s13071-015-1008-x

26. Li P, Xing H, Zhao Z, et al. Genetic diversity of Plasmodium falciparum histidine-rich protein 2 in the China-Myanmar border area. Acta Trop. 2015; 152: 26–31.

27. Kozycki CT, Umulisa N, Rulisa S, et al. False-negative malaria rapid diagnostic tests in Rwanda: impact of Plasmodium falciparum isolates lacking hrp2 and declining malaria transmission. Malar J. 2017; 16(1): 1–11.

28. Gendrot M, Fawaz R, Dormoi J, Madamet M, Pradines B. Genetic diversity and deletion of Plasmodium falciparum histidine-rich protein 2 and 3: a threat to diagnosis of P. falciparum malaria. Clin Microbiol Infect [Internet]. Elsevier Ltd; 2018;:2–7. Available from: https://doi.org/10.1016/j.cmi.2018.09.009

29. Ramutton. T, Hendriksen. IC, Mwanga-Amumpaire. J, et al. Sequence variation does not confound the measurement of plasma PfHRP2 concentration in African children presenting with severe malaria. Malar J [Internet]. 2012; 11:276. Available from: http://www.embase.com/search/results?subaction=viewrecord&from=export&id=L52169263%0Ahttp://dx.doi.org/10.1186/1475-2875-11-276%0Ahttp://sfx.library.uu.nl/utrecht?sid=EMBASE&issn=14752875&id=doi:10.1186%2F1475-2875-11-276&atitle=Sequence+variation+does+no

30. Thomson R, Beshir KB, Cunningham J, et al. Title:pfhrp2 and pfhrp3 gene deletions that affect malaria rapid diagnostic tests for for Plasmodium falciparum: analysis of archived blood samples from three African countries. 2019;.

31. Cheng Q, Gatton ML, Barnwell J, et al. Plasmodium falciparum parasites lacking histidine-rich protein 2 and 3: a review and recommendations for accurate reporting. Malar J [Internet]. 2014; 13(1):283. Available from: http://www.pubmedcentral.nih.gov/articlerender.fcgi?artid=4115471&tool=pmcentrez&rendertype=abstract

32. TACAIDS. Tanzania 2007–08 HIV/AIDS and Malaria Indicator Survey Key Findings. Tanzania 2007-08 HIV/AIDS Malar Indic Surv Key Find. 2008; (HIV Prevalence):16.

33. TACAIDS. Tanzania- 2011–12 HIV/AIDS and Malaria Indicator Survey 2011-12: Key Findings. Tanzania Comm AIDS (ZAC), Zanzibar AIDS Comm (NBS), Natl Bur Stat (OCGS), Off Chief Gov Stat ICF Int. 2013;:16.

34. TDHS-MIS. Tanzania Demographic and Health Survey and Malaria Indicator Survey (TDHS-MIS) 2015–16. Dar es Salaam, Tanzania, Rockville, Maryl USA [Internet]. 2016;:172-173. Available from: https://www.dhsprogram.com/pubs/pdf/FR321/FR321.pdf

35. THIS. Summary Sheet: Preliminary Findings 2016–2017. 2017; (December 2017):2016–2017. Available from: http://www.nbs.go.tz/nbs/takwimu/this2016-17/Tanzania_SummarySheet_English.pdf

36. Chacky F, Runge M, Rumisha SF, et al. Nationwide school malaria parasitaemia survey in public primary schools, the United Republic of Tanzania. Malar J [Internet]. BioMed Central; 2018; 17(1):1–16. Available from: https://doi.org/10.1186/s12936-018-2601-1

37. Chiduo MG, Mandara CI, Rumisha SF, et al. Assessing the intrinsic and extrinsic drivers and targeting the observed resilience of malaria in northwestern and southern Tanzania: A protocol for a cross-sectional exploratory study. 2020;:1–46.

38. MoH. National Guidelines for Diagnosis and Treatment of Malaria. Dar es Salam, Tanzania: Ministry of Health and Social Welfare, United Republic of Tanzania; 2006; (January).

39. Plucinski MM, Cruz KR, Ljolje D, et al. Screening for Pfhrp2/3 -Deleted Plasmodium falciparum, Non- falciparum, and Low-Density Malaria Infections by a Multiplex Antigen Assay. J Infect Dis. 2018; 219(3):437–447.

40. Lucchi NW, Karell MA, Journel I, et al. PET-PCR method for the molecular detection of malaria parasites in a national malaria surveillance study in Haiti, 2011. Malar J. 2014; 13:462.

41. Akerele D, Ljolje D, Talundzic E, Venkatachalam, Udhayakumar Lucchi WN. Molecular diagnosis of malaria by photo-induced electron transfer fluorogenic primers (PET-PCR). Am J Trop Med Hyg [Internet]. 2017; Conference:258. Available from: http://www.ajtmh.org/content/87/5_Suppl_1/226.full.pdf+html%5Cnhttp://ovidsp.ovid.com/ovidweb.cgi?T=JS&CSC=Y&NEWS=N&PAGE=fulltext&D=emed11&AN=71041482%5Cnhttp://eleanor.lib.gla.ac.uk:4550/resserv?sid=OVID:embase&id=pmid:&id=doi:&issn=0002-9637&isbn=&volum

42. Kudyba HM, Louzada J, Ljolje D, et al. Field evaluation of malaria malachite green loopmediated isothermal amplification in health posts in Roraima state, Brazil. Malar J [Internet]. BioMed Central; 2019; 18(1):1–7. Available from: https://doi.org/10.1186/s12936-019-2722-1

43. Sitali L, Miller JM, Mwenda MC, et al. Distribution of Plasmodium species and assessment of performance of diagnostic tools used during a malaria survey in Southern and Western Provinces of Zambia. Malar J [Internet]. BioMed Central; 2019; 18(1): 1–9. Available from: https://doi.org/10.1186/s12936-019-2766-2

44. Akinyi S, Magill AJ, Torres K, et al. Multiple genetic origins of histidine-rich protein 2 gene deletion in Plasmodium falciparum parasites from Peru. Sci Rep. 2013; 3(1): 1–8.

45. Abdallah JF, Okoth SA, Fontecha GA, et al. Prevalence of pfhrp2 and pfhrp3 gene deletions in Puerto Lempira, Honduras. Malar J. 2015; 14(1):1–9.

46. Plucinski M, Dimbu R, Candrinho B, et al. Malaria surveys using rapid diagnostic tests and validation of results using post hoc quantification of Plasmodium falciparum histidine-rich protein 2. Malar J. BioMed Central; 2017; 16(1): 1–7.

47. Benton CB, Nazha A, Pemmaraju N, et al. HHS Public Access. 2016; 95 (2): 222–242.

48. Luchivez J, Baker J, Alcantara S, et al. Laboratory demonstration of a prozone effect HRP2 detecting malaria diagnostic tests: implications for clinical management. Malar J [Internet]. 2011; 10:1–7. Available from: http://dx.doi.org/10.1186/1475-2875-10-286

49. Trouvay M, Palazon G, Berger F, et al. High Performance of Histidine-Rich Protein 2 Based Rapid Diagnostic Tests in French Guiana are Explained by the Absence of pfhrp2 Gene Deletion in P. falciparum. PLoS One. 2013; 8(9): 1–7.

